# Gait differences in patients with multiple sclerosis who have low and high levels of disability

**DOI:** 10.1101/19000166

**Authors:** John J. Fraser, Jeannie B. Stephenson

## Abstract

**Background:** Multiple Sclerosis (MS) often results in gait impairment and disability.

**Objective:** To investigate differences in spatiotemporal gait characteristics of people with MS who have low versus high levels of disability. Between trial and inter-limb consistency and the association of gait variables with level of disability were also investigated.

**Methods:** Participants with MS who had either low-disability [n=7; 3 females; EDSS mean: 2.7±0.5, range 2.0-4.5; BMI=26.9±6.6] or high-disability [n=11; 6 females; EDSS mean: 2.7±0.5, range 6.0-6.5; BMI=27.8±1.5) performed 2 trials of self-selected walking on an instrumented walkway. Differences in group, limb, and group by limb interactions were assessed using analysis of variance, independent-measures t-tests, and Cohen’s *d* effect sizes (ES). Between-trial consistency of gait were assessed with intra-class correlation coefficients (2, k).

**Results:** Participants in the high disability group had increased step time (ES=0.8), cycle time (ES=0.8), and ambulation time (ES=1.2) while taking shorter strides (ES=0.9) and more steps at a slower rate (ES=1.1). The high disability group demonstrated less between-trial consistency for 69% of gait variables when compared to the low disability group.

**Conclusion:** People with MS who have high levels of disability walk differently and with less consistency than those with lower levels of disability.

---

Multiple Sclerosis (MS) is a degenerative, autoimmune disorder that affects the central nervous system of approximately 400,000 individuals in the United States (Anderson et al., 1992) and usually manifests in the second to third decade of life.(Noseworthy et al., 2000) People with MS walk differently when compared to healthy individuals due to neurological impairment. They walk at slower velocities with an increased cadence, (Benedetti et al., 1999; Givon, Zeilig & Achiron, 2009; Sacco et al., 2011; Sosnoff et al., 2011; Sosnoff, Sandroff & Motl, 2012; Socie et al., 2013; Lizrova Preiningerova et al., 2015) demonstrate shorter step length,(Givon, Zeilig & Achiron, 2009; Sosnoff et al., 2011; Socie et al., 2013; Lizrova Preiningerova et al., 2015) and stride length,(Benedetti et al., 1999; Sacco et al., 2011) and a wider base of support.(Givon, Zeilig & Achiron, 2009; Sosnoff et al., 2011; Socie et al., 2013) People with MS also take longer to complete a single step (Givon, Zeilig & Achiron, 2009; Sosnoff et al., 2011; Socie et al., 2013; Lizrova Preiningerova et al., 2015) and to complete a gait cycle.(Benedetti et al., 1999; Sacco et al., 2011) They spend more time in periods of single(Sacco et al., 2011) and double limb support,(Benedetti et al., 1999; Sacco et al., 2011; Lizrova Preiningerova et al., 2015) and a decreased percentage of the gait cycle in swing phase.(Sosnoff et al., 2011) Sacco and colleagues (2011) also observed increased stride to stride variability in individuals with MS. Gait changes have been observed in individuals with MS who have varying degrees of impairment, with individuals with higher impairment demonstrating decreased gait velocity, step length, and step time and increased periods in double limb support.(Lizrova Preiningerova et al., 2015)

Clinically, the degree to which an individual is neurologically impaired and disabled is rated using the Expanded Disability Status Scale (EDSS).(Kurtzke, 1983) The EDSS is a ranking of disability ranging from 0 (normal) to 10 (death) determined by physical examination findings.(Kurtzke, 1983) The distance a person with MS can walk without rest breaks, support, or reliance on an assistive device (AD) are substantial factors in determination of the EDSS score.(Kurtzke, 1983) An EDSS score of 0 is representative of a normal neurological examination and full function. As the EDSS score increases to 5.5, the ability to walk distances without an AD is diminished to 100-m without rests or support. At an EDSS score of 6.0, the patient is dependent on a unilateral AD to walk 100-m without rests. While previous studies on the effect of AD use on spatiotemporal gait variables in participants with MS were performed using individuals who were relatively high functioning (median EDSS=4.8),(Gianfrancesco et al., 2011) further study is needed in participants with higher levels of disability. The purpose of this cross-sectional study was to investigate differences in spatiotemporal gait characteristics of people with MS who have low versus high levels of disability. Between trial and inter-limb consistency of these gait variables and the association of gait variables with level of disability were also investigated.

## METHODS

### Design

A cross-sectional, observational study was conducted using a sample of convenience of participants with MS in which the independent variables were group [mild to moderate (low) disability, high-disability] and lower limb (left, right) and the dependent variables were spatiotemporal gait parameters.

### Participants

Twenty-one participants diagnosed with MS were recruited from an MS Care Center. Sample size calculations estimated that 5 participants per group were needed to demonstrate large effect sizes for spatiotemporal gait variables in this population.(Benedetti et al., 1999) Prior to recruitment, a physical examination was performed by a board-certified neurologist who determined EDSS scores. Participants met inclusion criteria if they scored between 2.0 and 6.5 on the EDSS; demonstrated pyramidal or cerebellar symptoms on physical exam; and could ambulate at least 20 meters with or without an AD and/or orthotics. Group assignment was determined by EDSS score.(Kurtzke, 1983) Participants with mild to moderate disability (EDSS 2.0-4.5), who were able to walk 300-m without aid or rests, were assigned to the low-disability group.(Kurtzke, 1983) Participants with severe disability (EDSS 6.0-6.5), who relied on an AD to walk at least 20-meters, were assigned to the high-disability group.(Kurtzke, 1983) Participants were excluded if they were cognitively impaired and unable to follow instructions, pregnant, or had any medical condition that would substantially impair their ability to walk. Participants who met inclusion criteria provided informed consent and the study was approved by the Institutional Review Board at the College of Staten Island, City University of New York.

### Procedures

All data collection was performed mid-morning to mitigate the effects of fatigue commonly experienced by individuals with MS later in the day. Following enrollment, participants provided demographic information and height, mass and leg length were measured. Following familiarization with testing procedures, the subject walked the length of the instrumented walkway at a self-selected pace utilizing the AD they depended on for ambulation. Spatiotemporal gait parameters were collected using the GAITRite Walkway System (CIR Systems Inc., Clifton, NJ), a portable instrumented 8.3 m x 0.9 m mat which contains imbedded pressure-activated sensors and collects data at a sampling rate of 80 Hz. The GAITRite Walkway system has been shown to be both valid and reliable in the assessment of spatiotemporal gait parameters in adults with and without neurological deficit.(Nelson et al., 2002; Bilney, Morris & Webster, 2003; Menz et al., 2004) Two walking trials were performed at a self-selected pace separated by a 10-minute rest period.

### Data Processing

Spatiotemporal gait variables were processed using the GAITRite software package (CIR Systems Inc., Clifton, NJ). Prior to export of data from each trial, pressure readings introduced by an AD were deleted from each trial using the software footfall editor. The main outcome measures were spatial gait variables [Functional Ambulation Profile (FAP), a composite measure of gait performance(Gouelle, 2014)], gait velocity, cadence, step count, step length, stride length, width of base of support, toe out] and temporal gait variables (ambulation time, step time, cycle time, percent of gait cycle in swing and stance phase, stance time, and percentage of gait cycle in single and double limb support). Gait variable data represent the average of all footfalls across two walking trials.

### Statistical Analysis

Descriptive statistics were calculated for participant demographic and gait data. Group differences were assessed using the point estimate and 90% confidence intervals (CI) of the mean. Differences in group, limb, and group by limb interactions for gait variables that were limb specific (normalized step and stride length, base of support, toe out, step and cycle time, swing and stance %, and single and double limb support %) were assessed using a mixed factorial ANOVA. Independent measures t-tests were conducted for the dependent variables of FAP score, step count, cadence, normalized velocity, base of support, and ambulation time. Cohen’s *d* effect size (ES) point estimates and associated 90% CI were calculated to estimate the magnitude and precision of group differences. Effect sizes were interpreted ≥0.80 as large, 0.50-0.79 as moderate, 0.20-0.49 as small.(*Cohen, 2013*) Association of level of disability (AD use, EDSS score) to each gait variable was assessed using Spearman’s *ρ* correlation coefficient; interpreted ≥.75 as good to excellent, .50-74 moderate to good, .25-.49 fair, and 0-.25 little to no association.(*Portney & Watkins, 2009*) Consistency of gait trials for the low-disability and high-disability groups was assessed using the intraclass correlation coefficient (ICC, 2,*k*) and standard error of measurement. Data were analyzed using the Statistical Package for Social Sciences Version 23.0 (IBM, Inc., Armonk, New York, IL). The *a priori* level of statistical significance was set at *p* ≤0.05 for all analyses.

## RESULTS

The principle findings of this study were that there were differences in spatiotemporal gait variables in participants with MS who had high compared to low levels of disability and that there was an association between gait variables and level of disability.

Twenty-one adult participants with MS (10 males, 11 females) met criteria for inclusion in the study. Data from 18 participants (9 males, 9 females) were available for analysis. Participant demographic data are detailed in Table 1. One participant in the low-disability group and all participants in the high-disability group utilized an AD for walking trials. There were no significant differences between groups for age, BMI, or leg length. Gait data from three participants was corrupted and was excluded from analysis.

**TABLE 1.** Demographics of participants with Multiple Sclerosis with low or high-disability.

| | | Low-disability Group<br>(n=7, 4 males, 3 females)<br>Mean ± 90% CI | High-disability Group<br>(n=11, 5 males, 6 females)<br>Mean ± 90% CI |
| --- | --- | --- | --- |
| Age (years) | | 49.1±3.8 | 49.2±6.9 |
| BMI (kg/m<sup>2</sup>) | | 26.9±6.6 | 27.8±1.5 |
| Leg Length, (cm) | Lt | 90.4±3.1 | 90.8±2.5 |
| | Rt | 90.4±3.1 | 90.8±2.5 |
| Expanded Disability Severity Score | | 2.7±0.5 | 6.1±0.2 |
| Lt = left, Rt = right, cm=centimeters, SE=standard error, CI=confidence interval |  |  |  |

Group means, standard deviations, range, group main effect, limb main effect, group by limb interactions, and independent samples t-test results for spatiotemporal gait parameters are detailed in Table 2. Participants in the high disability group took significantly more steps (p=0.04) and required more time to walk along the 8.3-m walkway (p=0.02) than those in the low disability group. There were no other significant main effects for group, limb, or group by limb interactions.

**TABLE 2.**
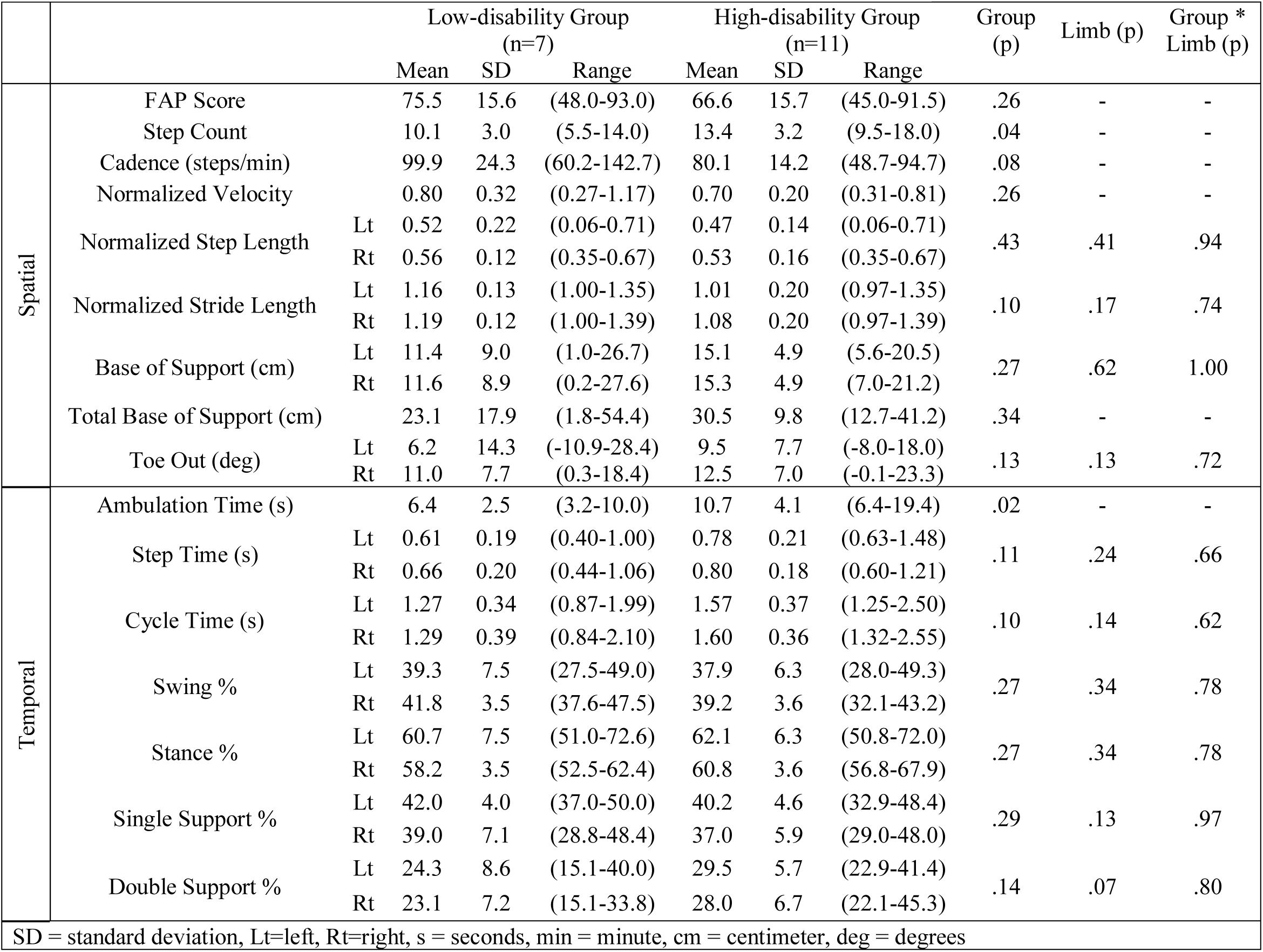
Gait variables in participants with Multiple Sclerosis with low or high-disability.

Cohen’s *d* ES point estimates and 90% CI comparing spatiotemporal gait variables in the high versus low disability group are illustrated in Figures 1 and 2. Participants with high disability took a longer time to complete a single step with the left foot (d=0.8), utilizing shorter stride lengths with the left leg (d=0.9). They took more steps (d=1.1) at a slower rate (d=1.1) and required more time to complete a gait cycle (Rt d=0.8, Lt d=0.9) and to walk the 8.3 m distance (d=1.0). While not statistically different, there was a trend toward decreased FAP scores (d=0.6), decreased normalized velocity (d=0.6), decreased normalized right stride length (d=0.7), decreased percentage of the gait cycle in swing phase (Rt d=0.7), increased right step time (d=0.8), increased percentage of the gait cycle spent in stance phase (Rt d=0.7), increased base of support (d=0.6), and increased periods of double limb support (Rt d=0.7, Lt d=0.8) observed in the high disability group (Figs. 1 & 2). These measures had moderate to large ES estimates, but with CI that crossed the ‘0’ threshold indicating statistical equivalence.

**FIGURE 1.**
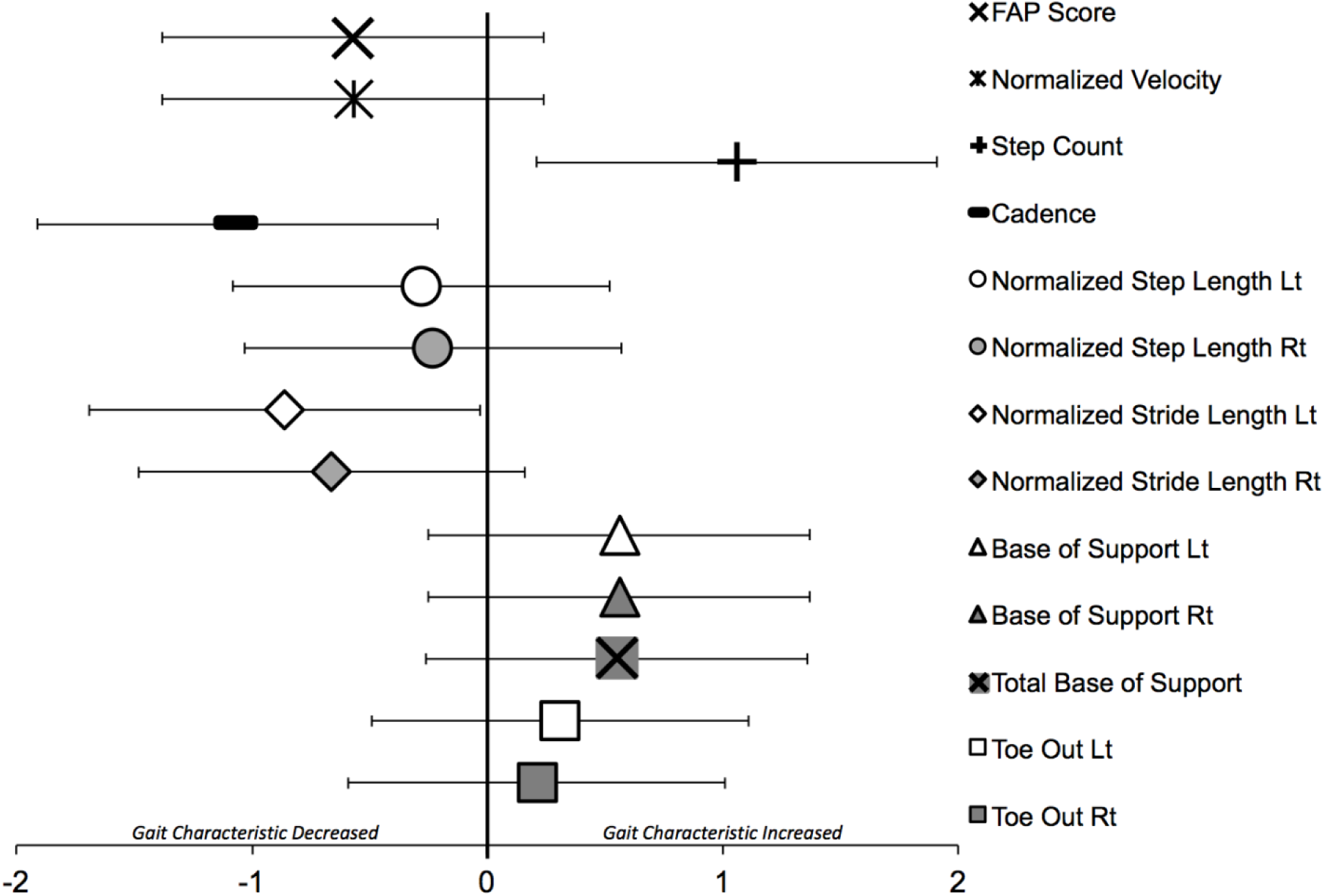
Cohen’s *d* effect size point estimates and 90% confidence intervals comparing spatial gait variables in participants with high-disability to participants with low-disability. FAP = Functional Ambulation Profile, Lt = left, Rt = right

**FIGURE 2.**
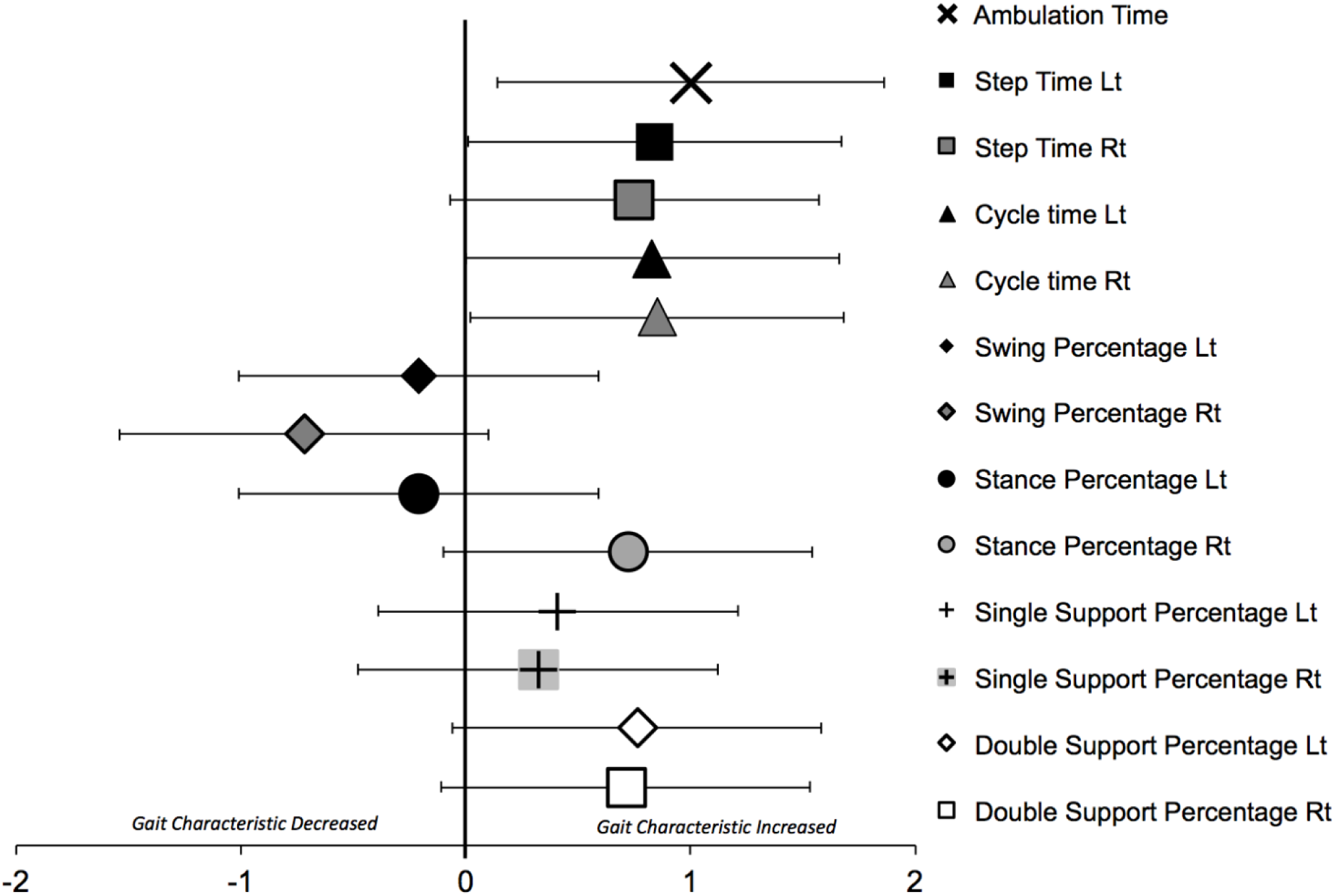
Cohen’s *d* effect size point estimates and 90% confidence intervals comparing temporal gait variables in participants with high-disability to participants with low-disability. FAP = Functional Ambulation Profile, Lt = left, Rt = right

In the low disability group, between-trial consistency was excellent for FAP scores, cadence, normalized velocity, limb dependent and total base of support, cycle time, and double limb support %, and fair to good for step count, ambulation time, and toe out (Table 3). The low disability group demonstrated variability between left and right lower limbs for normalized step and stride length, step time, swing %, stance %, and single limb support %. In the high-disability group, between-trial consistency was excellent for FAP scores, step count, and normalized stride length; fair to good for cadence, limb dependent and total base of support, toe out, ambulation time, step time, cycle time, swing %, and stance %; and poor for normalized step length. Inter-limb (left versus right) variability was noted in the high disability group for single limb support % and double limb support %.

**TABLE 3.**
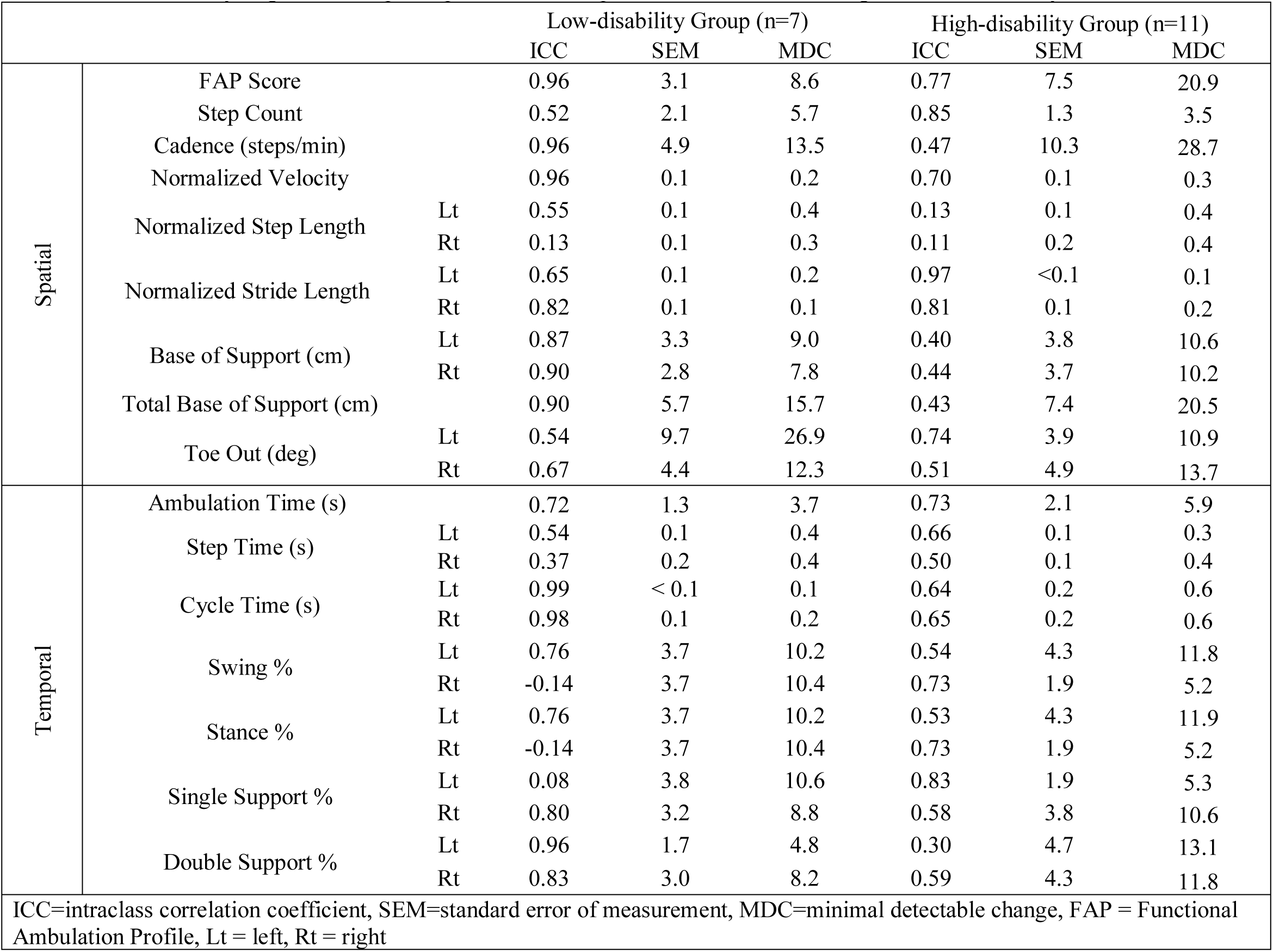
Consistency of gait trials in participants with Multiple Sclerosis with low or high levels of disability.

Associations between level of disability and gait variables are detailed in Table 4. Significant moderate to good associations were observed for use of an AD with step time, gait cycle time, and cadence. Association of higher disability, as measured by EDSS score, ranged from moderate to excellent for cadence, normalized stride length, ambulation time, step time, cycle time and percentage of time spent in double limb support.

**TABLE 4.**
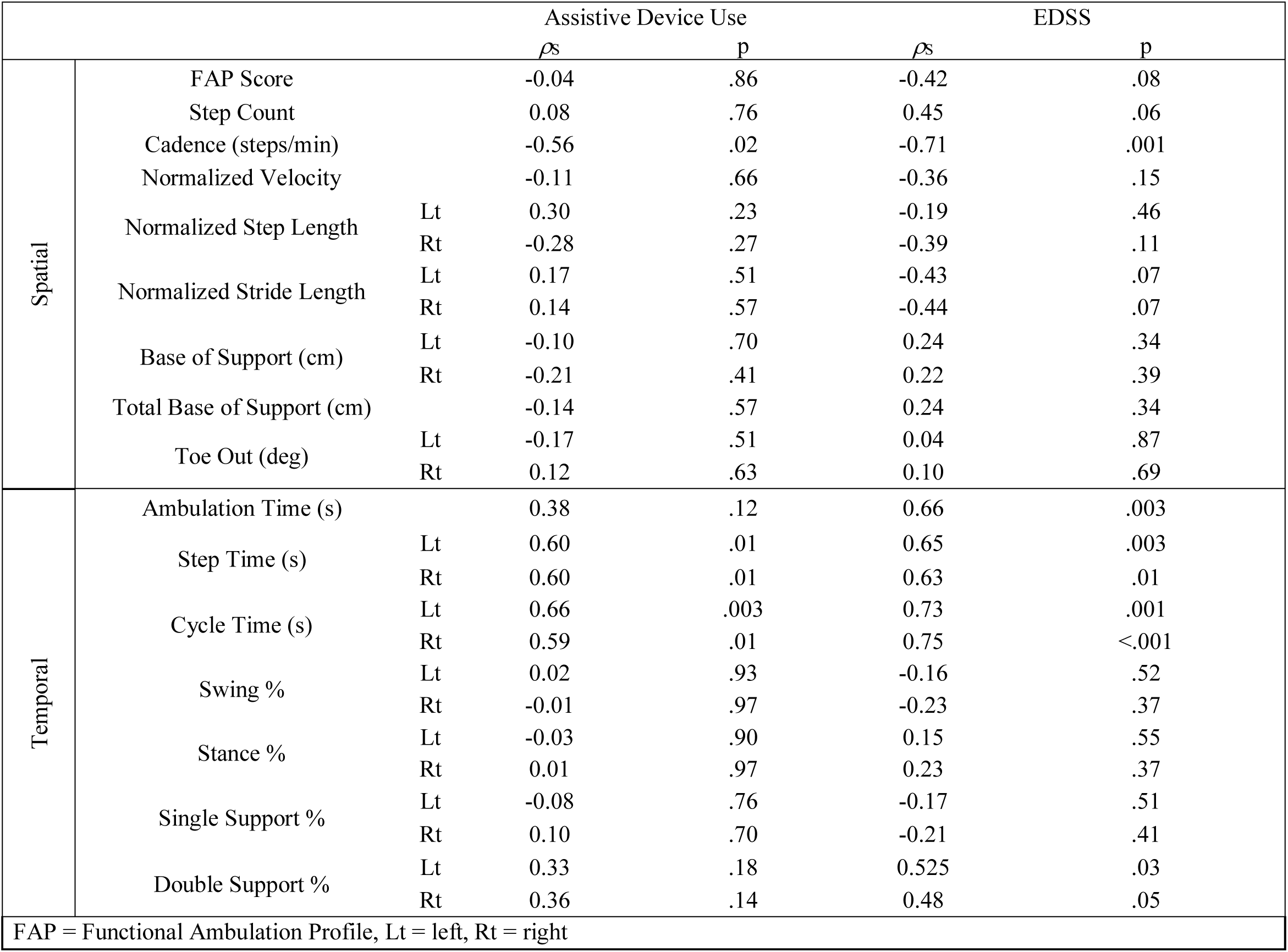
Association of gait variables with level of disability in participants with Multiple Sclerosis.

## DISCUSSION

To our knowledge, this is the first study to examine inter-limb spatiotemporal gait variables and consistency of gait in participants with MS with low versus high levels of disability.

### Gait differences

Our findings are consistent with results of previously published studies. Sosnoff and colleagues (2012) found that people with mild MS walked slower, with fewer, shorter and wider steps, and spent more time in double limb support than healthy controls. However, they did not compare these findings to participants with high disability.(Sosnoff, Sandroff & Motl, 2012) In another study, Sosnoff and colleagues (2011) found that MS participants with severe disability walked slower, took shorter steps and spent more time in double limb support when completing a cognitive task than participants with mild to moderate disability. Lizrova-Preiningerova and colleagues (2015) demonstrated that participants with MS with high disability walked slower and exhibited decreased step length, increased step time, and increased double limb support compared to individuals with MS with lower levels of disability. Although we did demonstrate a significant difference in step count and ambulation time between low and high disability groups in the present study, the differences found in gait velocity did not reach statistical significance. The larger sample size in the study conducted by Lizrova-Preiningerova and collegaues (Lizrova Preiningerova et al., 2015) may account for their finding of decreasing gait velocity with increasing EDSS scores.

The present study adds to these previous findings by also documenting inter-limb gait differences in individuals with low versus high levels of MS disability. Although there was no main effect for limb or group by limb interactions observed, there were significant differences noted in the ES for certain gait variables in one limb (shorter left stride length and longer left step time) and statistical equivalence in the contralateral limb. Wide CI observed in ES estimates may narrow with a larger sample size, demonstrating more distinct differences between groups for these gait variables. Knowledge of inter-limb differences in gait variables may be clinically meaningful information for physical therapists conducting gait training with these patients. For example, therapists could specifically address these inter-limb differences with therapeutic exercise or locomotor training to improve overall gait pattern consistency.

Previous studies have found that asymmetric lower extremity motor impairment of participants with MS adversely affects gait performance.(Chung et al., 2008; Sandroff, Sosnoff & Motl, 2013) High variability in many spatial and temporal gait parameters was observed in both disability groups in the present study. Socie and colleagues (2013) found decreased gait velocity, shorter step lengths, and increased step length variability in participants with MS who had higher disability compared to controls. Sosnoff and colleagues (2012) observed that participants with mild MS demonstrated greater step time variability and single support time variability than healthy, matched controls. Lizrova-Preiningerova and colleagues (2015) suggested that gait variability was not associated with level of disability in individuals with MS and may not be an appropriate outcome measure. However, the gait variability observed in the present study and in others indicates that this may be an appropriate measure of MS gait and an appropriate measure to use in intervention studies.(Sosnoff et al., 2011; Socie et al., 2013)

### Consistency of Gait Parameters between Trials and between Limbs

Both groups showed consistency of gait parameters between trials, but the low disability group was more consistent. Participants in the low disability group were more consistent in cadence, cycle time, base of support, percent of the gait cycle spent in stance and swing in the left leg, and in double limb support compared to participants in the high disability group. This is in contrast to the fair consistency observed in the high-disability group for % of the gait cycle spent in swing and stance phase. Socie and colleagues (2013) reported that participants with MS had greater variability in step and stride length compared to healthy controls and that variability increased with use of an AD. Inter-limb differences in consistency of normalized step length were observed in the present study in participants with high disability. This phenomenon was also observed for cadence, base of support, cycle time, and double limb support %, findings unique to this study. It is not surprising that individuals with higher levels of disability demonstrate less consistency and gait rehabilitation could be aimed at methods to specifically reduce between trial and inter-limb variability.

### Association of Gait Impairment and Disability

Assistive device use had a moderate to good association with cadence, step time and cycle time. Disability, as qualified by EDSS scores, had a strong association with gait cycle time and moderate to good association with cadence, ambulation time, step time, and double limb support %. Givon and colleagues (2009) found that EDSS score had a moderate to good association with FAP score, gait velocity, total base of support, and double limb support % in individuals with MS. Only the latter (double limb support %) was consistent with our results. Socie and colleagues (2013) found excellent association of EDSS score with gait velocity and moderate to good association with step length and step time. We demonstrated that those with higher disability took longer to walk than those with lower levels of disability, however, normalized velocity was not statistical significance. The disparate findings across these studies may be attributed to the heterogeneity of neurologic impairment unique to MS, which may manifest very differently between participants.

### Clinical Implications

Use of an instrumented walkway, in addition to observational gait analysis, may provide more objective information about spatiotemporal gait parameters in individuals with MS. Knowledge of MS gait patterns, particularly distinguishing those with low versus high disability, may allow physical therapists to tailor rehabilitation to patient’s individualized needs. Based on the findings of our study, clinicians evaluating patients with MS with higher levels of disability should consider assessing for and providing gait training to address changes in velocity, cadence, stride length, base of support, and proportion of time spent in stance and swing phase of gait. Assessment of consistency between walking trials and inter-limb consistency may also provide important insight into the patient’s ability to safely and efficiently execute the motor task of walking. Interventions to address spasticity and impaired lower extremity strength, power, endurance, and balance and providing appropriate assistive devices may help improve gait in these patients.(Lord, Wade & Halligan, 1998; Gutierrez et al., 2005; Chung et al., 2008; Cameron & Wagner, 2011; Sandroff, Sosnoff & Motl, 2013) Additionally, differences in gait parameters amongst individuals with low versus high MS disability should be considered when designing clinical trials.

### Study Limitations

The current study has some limitations. The authors relied on the EDSS score as the primary measure of disability, with additional delimitations of pyramidal or cerebellar deficit. Thus, participants assigned to each group were heterogeneous in presentation with possible differences in how they walked. This is evident in the wide CIs observed in the ES graphs in Figs. 1 and 2. For future research, we recommend that investigators who study individuals with MS with varying degrees of disability consider utilizing a patient reported outcome measure validated for use with this population, such as the Multiple Sclerosis Impact Scale, (Schäffler et al., 2013) to further qualify level of disability. Larger sample sizes and assessment of slow and fast walking speeds are required to demonstrate definitive group differences and provide further information about gait patterns in people with MS.

## CONCLUSIONS

Individuals with MS who have high levels of disability walk differently and with less consistency than those with lower levels of disability. Differences between left and right lower limbs were observed for certain gait variables and a high level of gait variability within groups and between gait trials were observed. Higher levels of disability were associated with shorter steps, decreased cadence, increased time in double limb support, greater gait variability and increased step time and gait cycle time. Clinically, it is important to delineate patients with MS who have low versus high levels of disability when examining gait and in order to provide tailored interventions to improve walking. More objective and longitudinal investigation of changes in gait characteristics of individuals with MS is needed to determine the most appropriate interventions at each stage of the disease.

## Data Availability

Data will be provided upon request.

## Acknowledgements

Cira Fraser, RN, PhD for her assistance in study design. Sanjeev Chopra, PT, MS, Sally Romano, PT, MS, and Luigi Leone PT, MS, and Aaron Miller, MD for their assistance in data collection. Jeffrey Rothman, PT, EdD for administrative support. Jay Hertel, PhD, ATC for his assistance in review and editing the manuscript. Arthur Nelson, PT, PhD^†^ for his guidance in conception, development, and oversight of the study. † Deceased 07 November 2010

## REFERENCES

Anderson DW, Ellenberg JH, Leventhal CM, Reingold SC, Rodriguez M, Silberberg DH. 1992. Revised estimate of the prevalence of multiple sclerosis in the United States. Annals of neurology 31:333–336.

Benedetti MG, Piperno R, Simoncini L, Bonato P, Tonini A, Giannini S. 1999. Gait abnormalities in minimally impaired multiple sclerosis patients. Multiple Sclerosis 5:363–368. DOI: 10.1177/135245859900500510.

Bilney B, Morris M, Webster K. 2003. Concurrent related validity of the GAITRite® walkway system for quantification of the spatial and temporal parameters of gait. Gait & Posture 17:68–74. DOI: 10.1016/S0966-6362(02)00053-X.

Cameron MH, Wagner JM. 2011. Gait abnormalities in multiple sclerosis: pathogenesis, evaluation, and advances in treatment. Current Neurology and Neuroscience Reports 11:507–515. DOI: 10.1007/s11910-011-0214-y.

Chung LH, Remelius JG, Van Emmerik REA, Kent-Braun JA. 2008. Leg power asymmetry and postural control in women with multiple sclerosis. Medicine and Science in Sports and Exercise 40:1717–1724. DOI: 10.1249/MSS.0b013e31817e32a3.

Cohen J. 2013. Statistical Power Analysis for the Behavioral Sciences. Academic Press.

Gianfrancesco MA, Triche EW, Fawcett JA, Labas MP, Patterson TS, Lo AC. 2011. Speed- and cane-related alterations in gait parameters in individuals with multiple sclerosis. Gait & Posture 33:140–142. DOI: 10.1016/j.gaitpost.2010.09.016.

Givon U, Zeilig G, Achiron A. 2009. Gait analysis in multiple sclerosis: Characterization of temporal–spatial parameters using GAITRite functional ambulation system. Gait & Posture 29:138–142. DOI: 10.1016/j.gaitpost.2008.07.011.

Gouelle A. 2014. Use of Functional Ambulation Performance Score as measurement of gait ability: Review. Journal of Rehabilitation Research and Development 51:665–674. DOI: 10.1682/JRRD.2013.09.0198.

Gutierrez GM, Chow JW, Tillman MD, McCoy SC, Castellano V, White LJ. 2005. Resistance Training Improves Gait Kinematics in Persons With Multiple Sclerosis. Archives of Physical Medicine and Rehabilitation 86:1824–1829. DOI: 10.1016/j.apmr.2005.04.008.

Kurtzke JF. 1983. Rating neurologic impairment in multiple sclerosis An expanded disability status scale (EDSS). Neurology 33:1444–1444. DOI: 10.1212/WNL.33.11.1444.

Lizrova Preiningerova J, Novotna K, Rusz J, Sucha L, Ruzicka E, Havrdova E. 2015. Spatial and temporal characteristics of gait as outcome measures in multiple sclerosis (EDSS 0 to 6.5). Journal of NeuroEngineering and Rehabilitation 12. DOI: 10.1186/s12984-015-0001-0.

Lord S, Wade D, Halligan P. 1998. A comparison of two physiotherapy treatment approaches to improve walking in multiple sclerosis: a pilot randomized controlled study. Clinical Rehabilitation 12:477–486.

Menz HB, Latt MD, Tiedemann A, Mun San Kwan M, Lord SR. 2004. Reliability of the GAITRite walkway system for the quantification of temporo-spatial parameters of gait in young and older people. Gait & Posture 20:20–25. DOI: 10.1016/S0966-6362(03)00068-7.

Nelson AJ, Zwick D, Brody S, Doran C, Pulver L, Rooz G, Sadownick M, Nelson R, Rothman J. 2002. The validity of the GaitRite and the Functional Ambulation Performance scoring system in the analysis of Parkinson gait. NeuroRehabilitation 17:255–262.

Noseworthy JH, Lucchinetti C, Rodriguez M, Weinshenker BG. 2000. Multiple Sclerosis. New England Journal of Medicine 343:938–952. DOI: 10.1056/NEJM200009283431307.

Portney LG, Watkins MP. 2009. Foundations of Clinical Research: Applications to Practice. Pearson/Prentice Hall.

Sacco R, Bussman R, Oesch P, Kesselring J, Beer S. 2011. Assessment of gait parameters and fatigue in MS patients during inpatient rehabilitation: a pilot trial. Journal of Neurology 258:889–894. DOI: 10.1007/s00415-010-5821-z.

Sandroff BM, Sosnoff JJ, Motl RW. 2013. Physical fitness, walking performance, and gait in multiple sclerosis. Journal of the Neurological Sciences 328:70–76. DOI: 10.1016/j.jns.2013.02.021.

Schäffler N, Schönberg P, Stephan J, Stellmann J-P, Gold SM, Heesen C. 2013. Comparison of patient-reported outcome measures in multiple sclerosis. Acta Neurologica Scandinavica 128:114–121. DOI: 10.1111/ane.12083.

Socie MJ, Motl RW, Pula JH, Sandroff BM, Sosnoff JJ. 2013. Gait variability and disability in multiple sclerosis. Gait & Posture 38:51–55. DOI: 10.1016/j.gaitpost.2012.10.012.

Sosnoff JJ, Boes MK, Sandroff BM, Socie MJ, Pula JH, Motl RW. 2011. Walking and Thinking in Persons With Multiple Sclerosis Who Vary in Disability. Archives of Physical Medicine and Rehabilitation 92:2028–2033. DOI: 10.1016/j.apmr.2011.07.004.

Sosnoff JJ, Sandroff BM, Motl RW. 2012. Quantifying gait abnormalities in persons with multiple sclerosis with minimal disability. Gait & Posture 36:154–156. DOI: 10.1016/j.gaitpost.2011.11.027.

